# Genetic characterization of a *Citrobacter braakii* isolate possessing *bla*_NDM-1_ obtained from toilet bowl water in a tertiary healthcare-associated facility in North Macedonia

**DOI:** 10.64898/2026.02.03.26345508

**Authors:** Nobuyoshi Yagi, Sora Miyazato, Fadil Cana, Ilir Demiri, Marija Cvetanovska, Georgi Eftimovski, Marija Dimzova, Itaru Hirai

## Abstract

Carbapenem-resistant (CR) bacteria have emerged and been spreading beyond healthcare-associated facilities into the environment. It is recognized that toilet bowl water in patient rooms of healthcare-associated facilities can be one of internal reservoirs of CR bacteria. In accordance with this idea, toilet bowl water samples were collected from patient rooms in a tertiary healthcare-associated facility in North Macedonia, and meropenem (MEM)-resistant bacterial isolates were obtained from the toilet bowl water. In this study, because a MEM-resistant *C. braakii* isolate, that was one of MEM-resistant opportunistic pathogens, was obtained from the toilet water, whole-genome sequencing (WGS) of this isolate was performed to obtain genetic characteristics of the *bla*_NDM-1_-positive *C. braakii* isolate. By the WGS, four contigs were constructed, the longest contig, contig 1 (5,189,681 bp), contained *bla*_CTX-M_ with some additional antimicrobial-resistance genes (ARGs). Interestingly, *bla*_NDM-1_ was detected in contig 2 (177,260 bp) and contig 3 (64,168 bp). Plasmid replicon of contig 2 was IncA/C2 but plasmid replicon of contig 3 was IncN and different from one of contig 2. Genetic structures surrounding *bla*_NDM-1_ were different between these two *bla*_NDM-1_-positive plasmids implying transfer or insertion of *bla*_NDM-1_ had occurred by IS or other mechanism. Further molecular epidemiology will be needed to explain the mechanism that allowed the *C. braakii* isolate to possess two structurally different *bla*_NDM-1_ plasmids.

## Introduction

*Citrobacter* species are Gram-negative bacilli that are widely distributed in the human and animal bodies, food items and the environment. Clinically, they are recognized as one of the causative agents of opportunistic infections. *C. freundii* has been frequently detected in clinical specimens. Even in the case of *C. freundii*, it still has not been easy to achieve species identification using classical yet golden standard biochemical tests that are utilized in automated microbial identification system in the clinical laboratories. Therefore, seven *Citrobacter* species, *i*.*e., C. braakii, C. freundii, C. gillenii, C. murliniae, C. rodentium, C. sedlakii* and *C. youngae* are handled as the *C. freundii* complex. In 1993, *C. farmer, C. youngae, C. braakii, C. werkmanii*, and *C. sedlakii* were named as new *Citrobacter* species based on the evaluation of genetic relatedness between strains using DNA hybridization technique [1] . However, there is not enough information regarding the detection rate of each *Citrobacter* species in medical settings, genetic characteristics of clinical *Citrobacter* isolates, and antimicrobial resistance profile of each *Citrobacter* species except for *C. freundii*.

*C. braakii* has been reported as a causative pathogen of diseases such as sepsis [2], peritonitis [3], bacteremia [4-6], and green nail syndrome [7]. The emergence and spread of antimicrobial-resistant (AMR) bacteria have become one of the major public health concerns, and the detection number of AMR *Citrobacter* species has also been increasing [8-10]. AMR *C. braakii* isolates have been isolated from many samples, such as river water [11-13], sewage [14, 15], food items [16-19], animal and human feces [20, 21], and patient specimens [22-26], and CR *C. braakii* strains have also been confirmed. It has been reported that *C. braakii* has acquired resistance to carbapenems through the production of OXA-48-type [12, 14, 24], VIM-type [13, 22, 24], NDM-type [14, 23, 25, 26], and KPC-type carbapenemases [13, 26].

Many studies analysis of CR Enterobacterales, such as *Escherichia coli* and *Acinetobacter baumannii*, indicated most of the carbapenemase genes were transferred among bacterial cells via transferrable plasmid [27-30]. However, genetic characteristics of plasmid found in *Citrobacter* spp. other than *C. freundii* have not been well characterized. Furthermore, it has been suggested that toilet bowl water in healthcare-associated facilities could be a new environmental reservoir for CR Enterobacteriaceae [31]. Consistent with this report, we isolated a CR *C. braakii* strain from the collected toilet bowl water of a tertiary healthcare-associated facility in North Macedonia. Therefore, in this study, we performed WGS of the isolated CR *C. braakii* isolate to clarify its genetic characteristics and to analyze the plasmid carrying *bla*_NDM-1_.

## Materials and Methods Isolation of CR bacteria

Seed swab kit (Eiken Chemical Co LTD., Tokyo, Japan) was used to collect toilet bowl water samples of toilets in two different patient rooms in a tertiary healthcare associated facility in North Macedonia. One toilet water sample was taken from each toilet, and in total two toilet water samples were collected. Cotton ball part of the seed swab kit was dipped in toilet bowl water and the wet cotton swab was inserted into Cary-Blair medium in the seed swab kit. The seed swab kits were transported to University of the Ryukyus, Japan and utilized to isolate antimicrobial-resistant bacteria.

To screen antimicrobial-resistance bacteria, the cotton ball parts of the seed swab kits ingrained the collected toilet water samples were used to streak them on MacConkey agar plates containing 2 μg/ml of cefotaxime (CTX) or 2 μg/ml of ciprofloxacin (CIP). After 24 hours at 37 °C, grown colonies were inoculated in Trypticase soy broth (BD Japan, Tokyo, Japan) containing CTX or CIP. Basically, three colonies were isolated from each sample. When differences in colony morphology or lactose fermentation were observed, three additional colonies representing each distinct phenotype were isolated for further analysis. The resulting bacterial stocks were kept at -80 °C until use. Susceptibility to meropenem (MEM) of the bacterial isolates was confirmed using Muller-Hinton (MH) agar containing μg/ml of MEM.

### WGS and data analysis

Basically, WGS of the CR *C. braakii* strain and data analysis using the obtained sequence information were performed as our previous study [32]. Briefly, extracted bacterial genomic DNA using the Monarch Spin gDNA Extraction Kit (New England Biolabs., Ipswich, MA) was subjected to Nanopore sequencing by using the Flongle Flow Cell (R10.4.1). The resulting nanopore sequencing reads were subjected to preferential deletion and quality trimming using Porechop [33] and NanoFilt [34]. Only reads with a quality score of 15 or higher and a read length of 2000 bp or longer were used for further analysis. The resulting draft genome was compared with reference genome sequences collected from RefSeq [35] using FastANI [36] for species identification, with an average nucleotide identity (ANI) cutoff of 95%. ARGs and plasmid replicons were detected using AMRFinderPlus [37] and PlasmidFinder [38], respectively. Sequence type (ST) of the sequenced *C. braakii* strain was determined at the PubMLST website [39]. To search for highly homologous plasmids, local BLAST analysis was performed on the plasmid sequences retrieved from the PLSDB [40]. Proksee was used to compare and visualize the database sequences with the *bla*_NDM-1_-positive plasmids derived from *C. braakii* in this study. DigAlign [41] was used to compare and visualize the surrounding genetic region of *bla*_NDM-1_ between the two *bla*_NDM-1_ positive plasmids confirmed in this study.

## Results

In this study, a total of 19 antibiotic-resistant bacteria were isolated from two toilet water samples, including 9 CTX-resistant strains and 10 CIP-resistant isolates. WGS was performed to identify bacterial species of the 19 AMR bacterial isolates. The results indicated the 19 AMR bacterial isolates included four isolates of *Acinetobacter baumannii*, three isolates each of *Aeromonas caviae, E. coli, Klebsiella oxytoca*, and *Pseudomonas aeruginosa* and one isolate each of *C. braakii, C. freundii*, and *Enterobacter asburiae*. Then, the bacterial isolates were inoculated on MH agar containing 1 ug/ml of MEM. Consequently, 13 of the 19 (68.4%) bacterial isolates were grown on MH agar containing MEM. Considering the MEM-resistance and the identified bacterial species, we selected one isolate, MEM-resistant *C. braakii*, for further genetic characterization.

By data analysis, four contigs were constructed from sequence reads obtained from MEM-resistant *C. braakii* isolate. ARGs were observed in all contigs (**Table 1** and **Supplementary data 1**), and there were major differences in composition and species of the detected ARGs. Among the detected ARGs, β-lactamase genes, such as *bla*_CTX-M_ and *bla*_NDM-1_ were confirmed in the contig 1, 2 and 3. Plasmid replicons were detected in the contig 2, 3 and 4, suggesting these three contigs were plasmids. ST was determined as ST281 by using the contig 1, implying contig 1 was chromosome. Collectively, the data indicated the MEM-resistant *C. braakii* isolates possessed two different *bla*_NDM-1_-harboring plasmids and one additional antimicrobial-resistance plasmid.

**Table 1.** Genetic chcracteristics of carbapenem-resistant *C. braakii* isolate.

| Contig # | length (bp) | ARGs | Replicon | ST |
| --- | --- | --- | --- | --- |
| 1 | 5,189,681 | **bla**<sub>CTX-M-15</sub>, *tet*(E), *tet*(E), *bla*<sub>CMY</sub>, *qnrB10* | - | 281 |
| 2 | 177,260 | *aph*(3')-VI, **bla**<sub>NDM-1</sub>, *sul1*, *armA*, *msr*(E),<br>*mph*(E), **bla**<sub>CMY-4</sub>, *dfrA14*, *arr-2*, *cmlA5*,<br>*bla*<sub>OXA-10</sub>, *aadA1*, *qacEdelta1*, *sul1* | IncA/C2 | - |
| 3 | 64,168 | *catB3*, **bla**<sub>NDM-1</sub>, *ble*, *dfrA25*, *qacEdelta1*,<br>*sul1*, *qnrB2*, *qacE*, *sul1* | IncN | - |
| 4 | 383,735 | *mph*(A), *sul1*, *qacEdelta1*, *catB3*, *bla*<sub>OXA-1</sub>,<br>*aac*(6')-Ib-cr5 | IncHI1A(CIT),<br>IncHI1B(CIT) | - |

Genetic similarity of the plasmids harboring *bla*_NDM-1_ to already published plasmids were considered with BLAST search, it indicated the contig 2 was similar to plasmids including *bla*_NDM-1_, such as **NZ_CP071074** (169, 082 bp), **MN657252.1** (168,682 bp) and **NZ_CP071071.1** (166,860 bp), and range of coverage of these plasmids were 95%, 94% and 94%, respectively (**Fig. 1A**). On the contrary, regarding the contig 3 (**Fig. 1B**), range of coverage rates was smaller than the case of the contig 2 and were 80 (**NZ_CP135452.1**, 85,586 bp) or less than 80 (**NZ_OQ821210.1**, 84,804 bp and **NZ_CP093467.1**, 58,125 bp), and interestingly, *bla*_NDM-1_ was not found in these three plasmids.

**Fig. 1.**
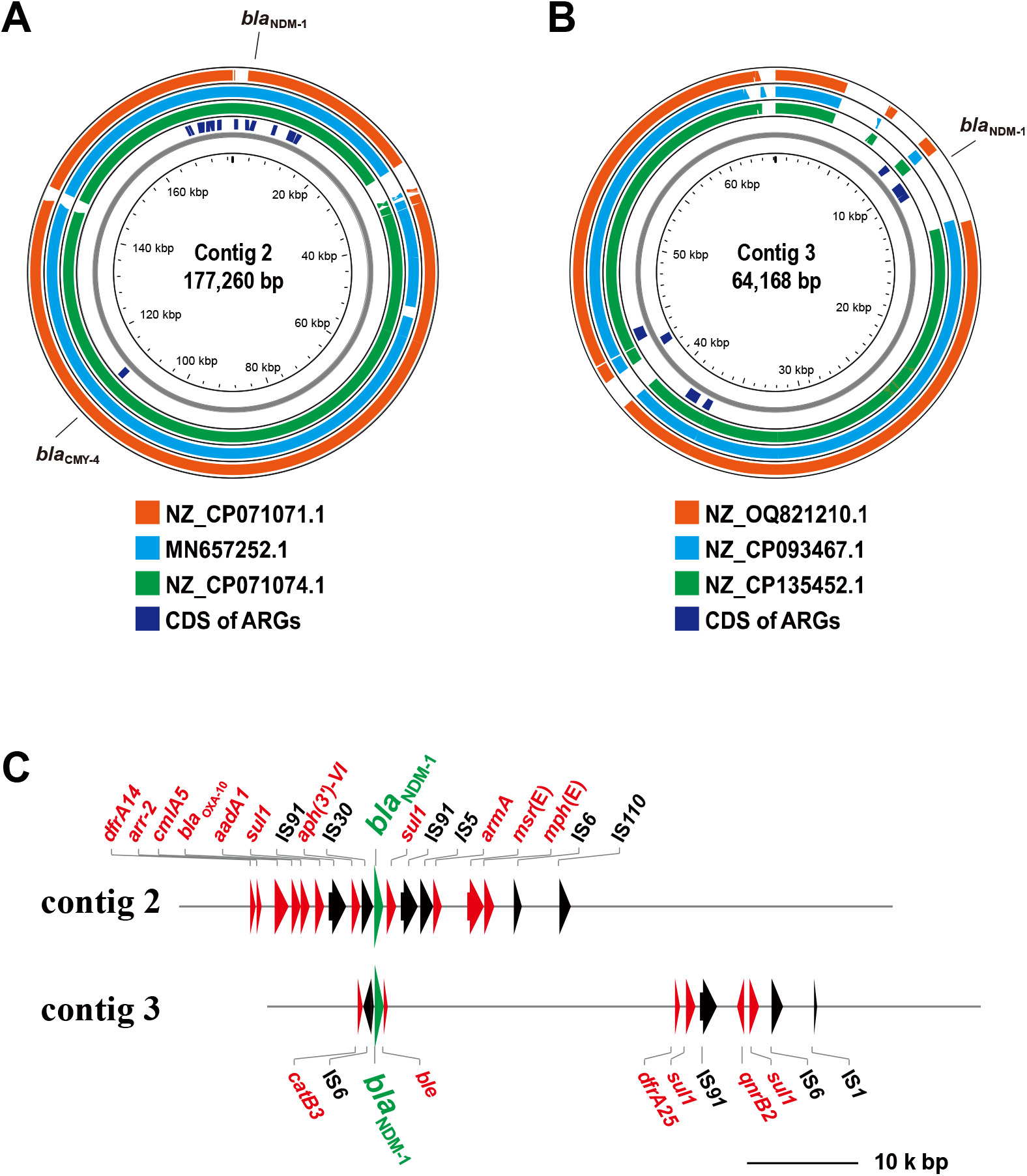
Similarity of *bla*_NDM-1_-positive plasmids detected in MEM-resistant *C. braakii* isolate. BLAST search was performed to retrieve similar plasmids to the two structurally different blaNDM-1-positive plasmids that were carried by *C. braakii* isolate. (**A**) Similarity of the IncA/C2 plasmid (contig 2). (**B**) Similarity of the IncN plasmid. Only detected beta-lactamase genes were indicated. (C) Genetic structures surrounding *bla*_NDM-1_ of the two *bla*_NDM-1_-positive plasmids are shown. Region surrounding *bla*_NDM-1_ of these two plasmids were extracted and depicted to compare whether there was any difference between these two regions.

In the MEM-resistant *C. braakii* isolates, one copy each of *bla*_NDM-1_ was found in the contig 2 and 3. To assess whether genetic structures surrounding *bla*_NDM-1_ were similar, region surrounding *bla*_NDM-1_ was extracted from the contig 2 and 3 and depicted. As shown in **Fig. 1C**, genetic structures surrounding *bla*_NDM-1_ were different between the contig 2 and 3, suggesting two different plasmids were introduced into the MEM-resistant *C. braaki*i strain rather than transfer of *bla*_NDM-1_ mediated by IS.

## Discussion

In this study, we genetically characterized a *bla*_NDM-1_-positive *C. braakii* strain isolated from the toilet bowl water in patient rooms of a tertiary healthcare-associated facility in North Macedonia. Our data interestingly showed that the *bla*_NDM-1_-positive *C. braakii* isolate simultaneously harbored two genetically distinct plasmids, such as IncA/C2 plasmid and IncN plasmid carrying *bla*_NDM-1_.

To date, *bla*_NDM-1_-positive IncA/C plasmids has been identified in Salmonella enterica isolated from migratory birds [42], variety of clinical isolates such as *Serratia marcescens* [43], *E. coli* in Myanmar [44], *E. coli* in Vietnam [45], *K. pneumoniae* in Vietnam [46], and *K. pneumoniae* in China [47]. However, similarity of genetic structures of these plasmids were not high enough to estimate distribution mechanisms of the *bla*_NDM-1_-positive IncA/C plasmid we detected in this study. As shown in **Fig, 1A**, BLAST analysis indicated the *bla*_NDM-1_-positive IncA/C plasmid was genetically similar to a *bla*_NDM-1_-positive plasmid (**MN657252.1**) carried by a *C. freundii* clinical isolate from a German hospital [48], and to plasmids carried by *C. sedlakii* (**NZ_CP071071.1**) or *E. coli* (**NZ_CP071074.1**) isolated in Switzerland by the GenBank genome information. From these results, it was possible that the *bla*_NDM-1_-positive IncA/C plasmid (contig 2) was introduced into North Macedonia via a European human, although it is unclear which bacterial strain(s) carried the IncA/C plasmid.

Regarding the *bla*_NDM-1_-positive IncN plasmid, there were reports from many regions such as Taiwan [49], China [50], Spain [51] and Ecuador [52]. However, our BLAST search showed the genetic similarity of the *bla*_NDM-1_-positive IncN plasmid was lower than the case of the contig 2. Three plasmids used in **Fig. 1B** did not contain *bla*_NDM-1_, implying insertion of DNA fragment including *bla*_NDM-1_ into conventional IncN plasmid.

In this study, we isolated the MEM-resistant and *bla*_NDM-1_-positive *C. braakii* isolate from toilet bowl water sample in a tertiary healthcare-associated facility in North Macedonia. Generally, it is regarded that *C. braakii* is not highly pathogenic to humans and can cause opportunistic infections. Therefore, it is important to note that the *bla*_NDM-1_ plasmid has been transmitted to such *C. braakii* and subsequently being excreted in toilets in patient rooms. Toilets or toilet bowl water have been proposed as a reservoir for CR bacteria within hospitals [31]. It is clear that the results obtained in this study provided another evidence supporting the idea.

The number of research reports on antimicrobial-resistant bacteria isolated in North Macedonia is not enough. In this regard, this study shed light on the nature of antimicrobial-resistant bacteria isolated in North Macedonia. However, at this point, the mechanism that allowed the MEM-resistant *C. braakii* isolate to harbor two structurally different *bla*_NDM-1_-positive plasmids is unknown. In addition to *bla*_NDM-1_, the MEM-resistant *C. braakii* isolate possessed multiple ARGs that conferred resistance to the same antimicrobial group, for instance, *bla*_CTX-M-15_ in chromosome (contig 1) and *bla*_CMY-4_ on the contig 2. These findings may reflect selective pressure from clinical antibiotic use. In any case, it is necessary to conduct more molecular epidemiological analysis targeting North Macedonia and collect more genome information of antimicrobial resistant bacteria to analyze the mechanism allowing the *C. braakii* isolate to possess the two structurally different *bla*_NDM-1_-positive plasmids.

## Data Availability Statement

Data available on request from the authors. The sequence reads used in this study are available under BioProject accession number **PRJDB37642**.

## Acknowledgements

This work was partly supported by the KAKENHI 23K09672 to IT and 24K20197 to NY.

## Notes

### Competing Interest Statement

The authors have declared no competing interest.

